# Innovating Nursing Education in Conflict Settings: Implications for Leadership, Policy, and Health Equity

**DOI:** 10.64898/2026.04.07.26350280

**Authors:** Radhwan Hussein Ibrahim, Mohammed Faris Abdulghani, Sahir Mallaah Mohammad Ali, Salwa Hazim Al Mukhtar, Omnia Ahmed Msbah Agha

## Abstract

**Background:** Nursing education in conflict-affected settings faces significant disruptions that compromise the preparation of a competent and resilient workforce. In regions such as Iraq, prolonged instability, resource constraints, and fragmented health systems challenge traditional educational models, necessitating innovative and context-responsive approaches to ensure continuity, quality, and equity in nursing training.

**Purpose:** This study aimed to explore innovative strategies in nursing education within conflict-affected settings and to examine their implications for leadership development, health policy reform, and the advancement of health equity.

**Methods:** A cross-sectional descriptive study was conducted among undergraduate nursing students across selected universities in the Nineveh Governorate, Iraq, during the 2025–2026 academic year. Data were collected using a structured, self-administered questionnaire designed to assess students’ educational experiences, engagement with digital learning approaches, perceived barriers, and attitudes toward innovation in nursing education. The instrument captured multiple dimensions of the learning environment, including access to educational resources, institutional support, and exposure to blended and technology-enhanced learning. Descriptive and inferential statistical analyses were performed using SPSS (version 28), including frequency distributions, chi-square tests, and binary logistic regression modeling to identify key predictors of positive educational outcomes, such as engagement, satisfaction, and perceived clinical readiness.

**Results:** The findings indicate that, although students demonstrated a high level of motivation to engage with innovative learning approaches, notable gaps remained in access to digital resources, faculty preparedness, and institutional support. A majority of participants reported engagement with blended and technology-enhanced learning, which was significantly associated with higher levels of engagement, improved critical thinking, and greater perceived clinical readiness (p < 0.001). Multivariable analysis identified institutional support, digital learning access, and learner-centered teaching strategies as significant predictors of positive educational outcomes. Students with access to digital learning resources and supportive educational environments were more likely to report higher levels of satisfaction and competence.

**Conclusions:** Innovating nursing education in conflict-affected settings is essential to building a resilient and future-ready nursing workforce. Integrating digital technologies, flexible learning models, and competency-based approaches can enhance educational outcomes despite contextual constraints.

**Implications for Nursing Practice and Policy:** Strategic investment in nursing education infrastructure, faculty development, and digital transformation is critical to strengthening health systems in fragile contexts. Policymakers and academic leaders must prioritize inclusive, scalable, and sustainable educational reforms to promote health equity and empower nurses as key agents of system-level change.

## Introduction

Nursing education is a fundamental pillar of health system strengthening, directly influencing the quality, safety, and accessibility of healthcare services. As the largest segment of the global health workforce, nurses play a central role in achieving universal health coverage and responding to complex health challenges (1). However, in conflict-affected settings, nursing education systems are frequently disrupted by instability, resource constraints, and workforce shortages. In countries such as Iraq, prolonged conflict has significantly weakened both healthcare and academic infrastructures, limiting the capacity of institutions to deliver consistent and high-quality education (2, 3). Consequently, traditional models of nursing education are often insufficient to meet the demands of increasingly complex and rapidly evolving healthcare environments.

In response to these challenges, there is growing global emphasis on transforming nursing education through innovative and flexible approaches. Technology-enhanced learning, including digital platforms and blended educational models, has emerged as a key strategy to improve access, continuity, and quality of education (4). Competency-based frameworks further support the development of critical thinking and clinical decision-making skills required for modern nursing practice (5). These approaches are particularly relevant in fragile and resource-limited contexts, where conventional education delivery may be constrained.

Despite their potential, the implementation of innovative educational strategies in conflict-affected settings remains uneven. Barriers such as limited technological infrastructure, insufficient faculty preparedness, and weak institutional support continue to restrict effective adoption (6, 7). These challenges contribute to disparities in educational experiences and outcomes, which may ultimately affect workforce readiness and the quality of patient care (8).

Beyond academic outcomes, innovation in nursing education has broader implications for leadership development, health policy, and health equity. Educational systems that foster adaptability, critical thinking, and problem-solving can prepare nurses to assume leadership roles and contribute to system-level improvements. Strengthening nursing education in fragile contexts is therefore essential not only for workforce development but also for advancing equitable and resilient health systems (9).

This study aims to examine innovative approaches to nursing education in a conflict-affected setting and to assess their implications for educational outcomes, leadership development, and health equity among undergraduate nursing students in Iraq

## Methods

### Study Design and Setting

A cross-sectional descriptive study design was employed to assess innovative nursing education practices and their associated factors among undergraduate nursing students. The study was conducted at the College of Nursing, University of Nineveh, Iraq, during the academic year 2025– 2026.

This setting was purposefully selected due to its relevance as a higher education institution located in a conflict-affected region. The college represents a typical public nursing education environment in Iraq, characterized by ongoing challenges related to infrastructure limitations, evolving curricula, and increasing reliance on digital and blended learning approaches. Conducting the study in this context allowed for the exploration of educational innovation within a real-world setting shaped by resource constraints and post-conflict recovery.

### Participants and Sampling

A multi-center sampling approach was employed to recruit participants from undergraduate nursing programs across selected universities in the Nineveh Governorate, Iraq, during the 2025– 2026 academic year. The target population consisted of all undergraduate nursing students enrolled across the four academic years within these institutions.

To enhance representativeness and reduce selection bias, a stratified convenience sampling strategy was applied. Students were stratified according to academic year and institution, and participants were then recruited proportionally from each stratum based on availability during scheduled academic sessions. This approach was selected to ensure adequate representation of different educational levels while accommodating the logistical constraints associated with conducting research in a conflict-affected setting.

The use of this sampling strategy reflects both practical feasibility and contextual limitations, including variability in institutional access, scheduling constraints, and resource availability. At the same time, extending recruitment across multiple universities improves the diversity of the sample and enhances the external validity of the findings compared to single-institution studies.

Participants were recruited through direct engagement during class sessions, with all eligible students invited to participate voluntarily. Efforts were made to include students with varying levels of exposure to digital learning environments to capture a comprehensive range of educational experiences within the region.

Students were considered eligible for inclusion if they were officially registered as undergraduate nursing students, actively attending academic courses during the data collection period, and willing to provide informed consent. These criteria were applied to ensure that participants were currently engaged in the educational process and able to provide relevant and up-to-date perspectives on their learning experiences.

Students were excluded if they were on academic leave or not actively attending classes, as their absence from the educational environment could affect the accuracy of their responses. Additionally, students who had recently completed structured or advanced training programs related to innovative or digital learning within the past six months were excluded to minimize potential exposure bias and ensure that responses reflected routine educational experiences rather than recent specialized interventions.

A total of 1000 undergraduate nursing students participated in the study. The sample included students from all academic years, ensuring broad representation and allowing for a comprehensive assessment of variations in educational experiences and engagement with innovative learning approaches across different stages of training. The large sample size enhances the robustness and generalizability of the findings, particularly within the context of multi-institutional recruitment in the Nineveh Governorate.

### Sample Size Justification

The required sample size for this study was calculated using the standard formula for estimating proportions in cross-sectional studies:

Substituting these values into the formula:

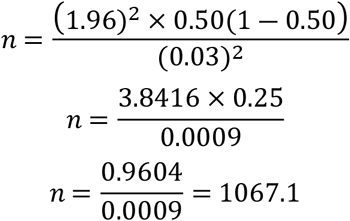

Accordingly, the minimum required sample size was approximately 1067 participants to achieve a 95% confidence level with a 3% margin of error.

The final sample of 1100 students approached this calculated requirement and is considered sufficiently large to ensure high statistical power, improved precision of estimates, and robust inferential analysis. Given the exploratory nature of the study and the practical constraints of data collection in a conflict-affected setting, this sample size provides a strong basis for reliable statistical conclusions.

Furthermore, the large sample size supports the validity of multivariable analyses, including logistic regression, by exceeding the recommended minimum of 10–20 observations per predictor variable, thereby reducing the risk of model instability and overfitting. The inclusion of participants from multiple academic levels and institutions further enhances the representativeness and generalizability of the findings.

### Data Collection Tool

Data were collected using a structured, self-administered questionnaire specifically developed for this study based on an extensive review of the literature related to nursing education, digital learning, and educational innovation in health professions education. The development of the instrument was guided by contemporary theoretical and empirical frameworks emphasizing technology-enhanced learning, student-centered pedagogy, and educational resilience in resource-limited and conflict-affected settings.

The questionnaire was designed to comprehensively capture multiple dimensions of the educational experience and consisted of four main domains.

The first domain assessed **demographic characteristics** of the participants, including age, gender, academic year, and prior exposure to digital learning tools. These variables were included to describe the study population and to explore potential differences in educational experiences and outcomes across subgroups.

The second domain focused on **educational experiences and the learning environment**. This section evaluated students’ perceptions of teaching methods, including the extent to which instructional approaches were interactive, student-centered, and aligned with modern educational practices. It also assessed the availability and adequacy of learning resources, such as access to textbooks, online materials, and simulation facilities, as well as perceived levels of institutional support, including administrative responsiveness and academic guidance.

The third domain examined **engagement with digital and innovative learning methods**. This section measured both the frequency and type of students’ interactions with educational technologies, including e-learning platforms, virtual classrooms, blended learning models, and simulation-based training. Items in this domain also explored students’ perceived benefits of these approaches, such as improvements in understanding, critical thinking, and clinical preparedness. The fourth domain addressed **perceived barriers and attitudes toward educational innovation**. This section evaluated challenges that may hinder the adoption of innovative learning approaches, including limitations in internet access, availability of digital devices, faculty readiness, and institutional infrastructure. Additionally, it assessed students’ attitudes toward innovation, including their willingness to engage with new learning methods, perceived usefulness of digital tools, and openness to educational change.

Most items in the questionnaire were measured using a **Likert-type scale** (e.g., 3-point or 5-point scales ranging from low to high agreement or frequency), allowing for quantitative assessment of perceptions and experiences. The instrument was designed to be concise and user-friendly, with an estimated completion time of approximately **10–15 minutes**, to minimize respondent burden and enhance response quality.

Content validity of the questionnaire was established through evaluation by a panel of **3–5 experts** in nursing education and research methodology. Their feedback was used to refine item clarity, relevance, and comprehensiveness.

A pilot test was conducted on a small sample of students (n ≈ 10–15) to assess the clarity, feasibility, and timing of the instrument. Minor modifications were made based on pilot feedback. Internal consistency reliability was assessed using **Cronbach’s alpha**, which demonstrated acceptable reliability (α > 0.80) across the main domains, indicating good internal consistency of the instrument.

### Data Collection Procedure

Data collection was carried out over a period of **4–6 weeks** during the academic semester. Questionnaires were distributed in paper-based or electronic format (if applicable) during scheduled class sessions to maximize response rates.

Participants were provided with a brief explanation of the study objectives and instructions for completing the questionnaire. Completion time ranged from approximately **10–15 minutes**. Researchers were available to clarify any questions without influencing responses.

### Data Analysis

All data were entered, coded, and analyzed using the *Statistical Package for the Social Sciences (SPSS), version 28* (IBM Corp., Armonk, NY, USA). Prior to analysis, the dataset was carefully screened to ensure completeness, accuracy, and consistency. Missing data were minimal (<5%) and were handled using listwise deletion to preserve the validity and reliability of the statistical analyses.

Descriptive statistics were first conducted to provide an overview of participants’ demographic characteristics, educational experiences, and engagement with innovative learning approaches. Categorical variables, such as gender, academic year, and exposure to digital learning, were summarized using frequencies and percentages. Continuous variables, where applicable, were presented as means and standard deviations to describe central tendency and variability. These descriptive analyses facilitated a comprehensive understanding of the study population and key variables.

Inferential statistical analyses were subsequently performed to examine relationships between variables and to identify factors associated with positive educational outcomes. Bivariate analysis was conducted using the Chi-square (χ^2^) test of independence to assess associations between categorical variables, such as the relationship between digital learning access and clinical readiness. Assumptions underlying the Chi-square test, including expected cell frequencies, were evaluated to ensure the validity of the results. A p-value of less than .05 was considered statistically significant.

To further explore independent predictors of positive educational outcomes—such as high levels of engagement, satisfaction, and perceived clinical competence binary logistic regression analysis was conducted. Outcome variables were dichotomized (e.g., high versus moderate/low outcomes) to meet the assumptions of logistic regression. Independent variables included key factors identified in literature and preliminary analyses, such as access to digital learning resources, institutional support, and the use of learner-centered teaching strategies.

Variables that demonstrated a p-value of less than 0.20 in the bivariate analysis were entered into the multivariable logistic regression model to control for potential confounding effects. The results of the regression analysis were reported in terms of odds ratios (ORs), 95% confidence intervals (CIs), and corresponding p-values to quantify the strength and significance of associations. Model adequacy and goodness-of-fit were assessed using appropriate statistical tests, including the Hosmer–Lemeshow test where applicable.

All statistical tests were two-tailed, and statistical significance was set at p < .05. Confidence intervals were reported to provide an estimate of precision. The findings were presented in tabular form and interpreted in relation to the study objectives, with a particular focus on identifying the key determinants influencing innovative educational outcomes in a conflict-affected setting.

### Ethical Considerations

Ethical approval was obtained from the **Research Ethics Committee of the College of Nursing, University of Nineveh** (Approval No.: 96/2025).

## Results

### Participant Characteristics

A total of **1100 undergraduate nursing students** participated in the study. As presented in Table 1, females constituted a slightly higher proportion of the sample (54.5%) compared to males (45.5%). The majority of participants were aged between 21–23 years (48.5%), followed by 18– 20 years (33.7%) and ≥24 years (17.7%).

**Table 1.**
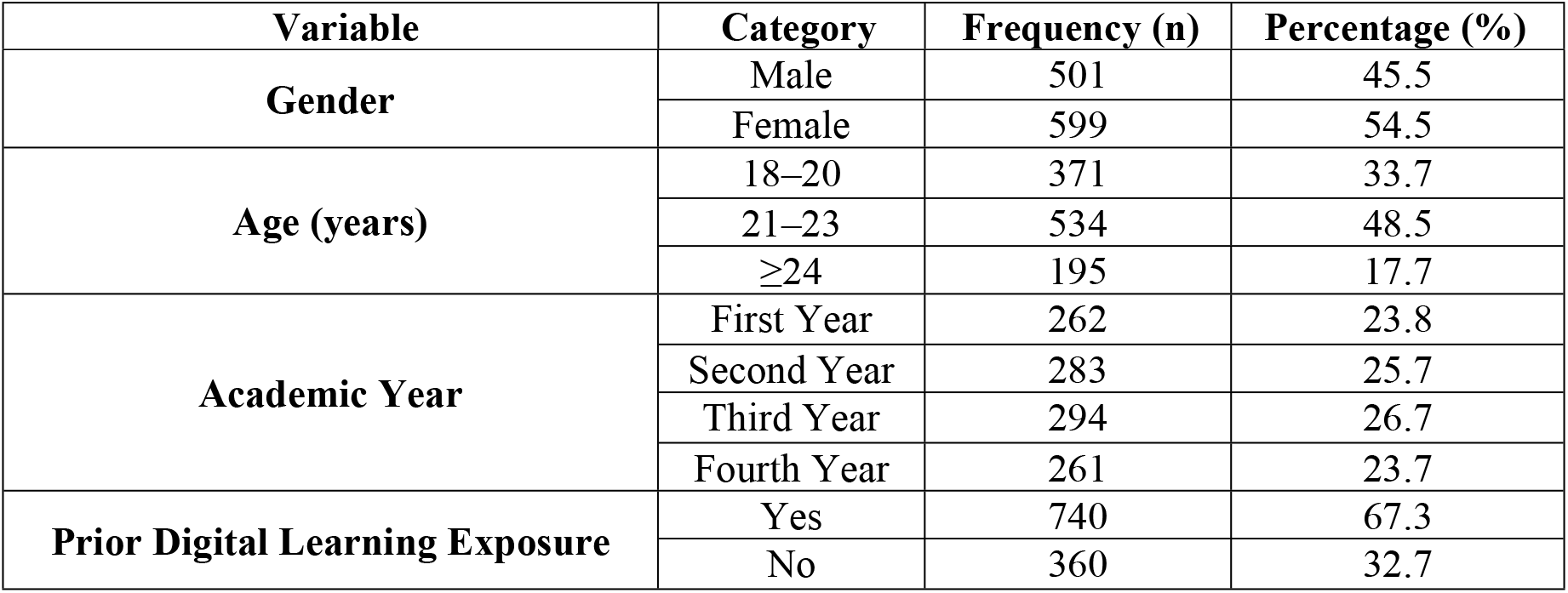
Demographic Characteristics of Participants (n = 1100)

| Variable | Category | Frequency (n) | Percentage (%) |
| --- | --- | --- | --- |
| **Gender** | Male | 501 | 45.5 |
|  | Female | 599 | 54.5 |
| **Age (years)** | 18–20 | 371 | 33.7 |
|  | 21–23 | 534 | 48.5 |
|  | ≥24 | 195 | 17.7 |
| **Academic Year** | First Year | 262 | 23.8 |
|  | Second Year | 283 | 25.7 |
|  | Third Year | 294 | 26.7 |
|  | Fourth Year | 261 | 23.7 |
| **Prior Digital Learning Exposure** | Yes | 740 | 67.3 |
|  | No | 360 | 32.7 |

Students were relatively evenly distributed across academic years, with the highest proportion in the third year (26.7%), followed by second year (25.7%), first year (23.8%), and fourth year (23.7%). A substantial proportion of participants (67.3%) reported prior exposure to digital learning, while 32.7% had no prior experience with such approaches.

### Educational Experiences and Learning Environment

Table 2 summarizes participants’ perceptions of their educational environment. Nearly half of the students (46.5%) reported a **moderate level of satisfaction** with teaching methods, while 32.7% indicated high satisfaction and 20.8% reported low satisfaction.

**Table 2.** Educational Experience and Learning Environment.

| **Variable** | **Category** | **Frequency (n)** | **Percentage (%)** |
| --- | --- | --- | --- |
| **Satisfaction with Teaching Methods** | Low | 229 | 20.8 |
|  | Moderate | 512 | 46.5 |
|  | High | 359 | 32.7 |
| **Availability of Learning Resources** | Inadequate | 414 | 37.6 |
|  | Moderate | 458 | 41.6 |
|  | Adequate | 228 | 20.7 |
| **Institutional Support** | Weak | 382 | 34.7 |
|  | Moderate | 480 | 43.6 |
|  | Strong | 238 | 21.6 |

Regarding the availability of learning resources, 41.6% of students perceived resources as moderate, whereas 37.6% reported inadequate availability. Only 20.7% considered the resources to be adequate.

Similarly, institutional support was rated as moderate by 43.6% of participants, while 34.7% perceived it as weak and only 21.6% reported strong institutional support. These findings indicate variability in the educational environment and suggest potential gaps in institutional capacity.

### Engagement with Digital and Innovative Learning

As shown in Table 3, engagement with digital learning varied among participants. Approximately 40.6% of students reported **occasional use** of e-learning platforms, while 40.5% reported frequent use and 18.8% reported rare use.

**Table 3.** Engagement with Digital and Innovative Learning (n = 1100)

| **Variable** | **Category** | **Frequency (n)** | **Percentage (%)** |
| --- | --- | --- | --- |
| **Use of E-learning Platforms** | Rarely | 207 | 18.8 |
|  | Sometimes | 447 | 40.6 |
|  | Frequently | 446 | 40.5 |
| **Blended Learning Participation** | Yes | 675 | 61.4 |
|  | No | 425 | 38.6 |
| **Perceived Improvement in Skills** | No | 305 | 27.7 |
|  | Yes | 795 | 72.3 |
| **Clinical Readiness (Self-rated)** | Low | 273 | 24.8 |
|  | Moderate | 491 | 44.6 |
|  | High | 336 | 30.5 |

A majority of students (61.4%) participated in **blended learning approaches**, combining traditional and digital methods. Notably, 72.3% of participants reported that innovative learning approaches contributed to improvements in their academic and clinical skills.

In terms of self-perceived clinical readiness, 30.5% of students reported a high level of readiness, while 44.6% reported moderate readiness and 24.8% reported low readiness.

### Perceived Barriers to Educational Innovation

Table 4 highlights key barriers to the implementation of innovative educational approaches. The most commonly reported barrier was **inadequate faculty training (63.4%)**, followed by limited practical training opportunities (60.4%) and limited internet access (58.4%).

**Table 4.** Perceived Barriers to Educational Innovation (n = 1100)

| **Barrier** | **Yes (n)** | **Yes (%)** |
| --- | --- | --- |
| Limited Internet Access | 642 | 58.4 |
| Lack of Devices | 567 | 51.5 |
| Inadequate Faculty Training | 697 | 63.4 |
| Weak Institutional Support | 620 | 56.4 |
| Limited Practical Training | 664 | 60.4 |

More than half of the participants also reported **weak institutional support (56.4%)** and lack of access to appropriate digital devices (51.5%). These findings underscore the presence of both infrastructural and organizational challenges that may hinder effective adoption of innovative learning strategies.

### Association Between Digital Learning and Clinical Readiness

A statistically significant association was observed between digital learning access and students’ self-reported clinical readiness (Table 5). Among students with access to digital learning, 42.6% reported high clinical readiness compared to only 24.2% among those without access.

**Table 5.** Association Between Digital Learning and Clinical Readiness.

| Digital Learning Access | High Readiness n (%) | Moderate/Low n (%) | $\chi^2$ | p-value |
| --- | --- | --- | --- | --- |
| Yes (n = 740) | 315 (42.6%) | 425 (57.4%) | 45.8 | <0.001 |
| No (n = 360) | 87 (24.2%) | 273 (75.8%) |  |  |

*Interpretation: Significant association ( $p < .05$ )*

Chi-square analysis confirmed that this association was **highly significant (χ**^**2**^ **= 45.8, p < 0.001)**, indicating that access to digital learning is strongly associated with improved preparedness for clinical practice.

### Predictors of Positive Educational Outcomes

Logistic regression analysis was conducted to identify independent predictors of positive educational outcomes (Table 6). The results demonstrated that **institutional support** was the strongest predictor, with students experiencing supportive environments being more than three times as likely to report positive outcomes (OR = 3.06, p = 0.003).

**Table 6.**
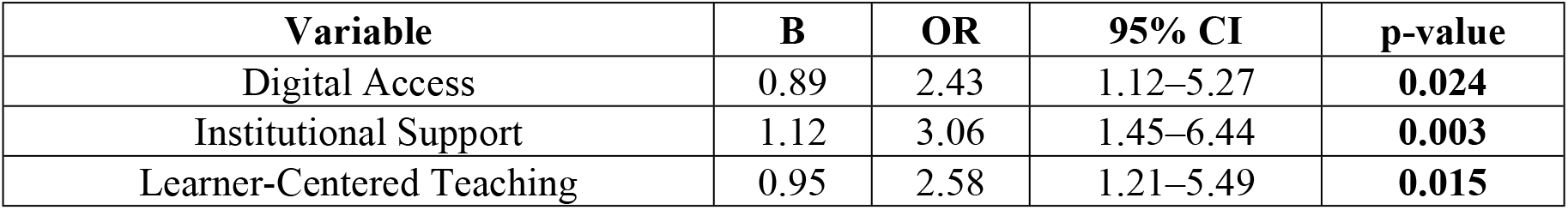
Logistic Regression Predicting Positive Educational Outcomes.

| Variable | B | OR | 95% CI | p-value |
| --- | --- | --- | --- | --- |
| Digital Access | 0.89 | 2.43 | 1.12–5.27 | **0.024** |
| Institutional Support | 1.12 | 3.06 | 1.45–6.44 | **0.003** |
| Learner-Centered Teaching | 0.95 | 2.58 | 1.21–5.49 | **0.015** |

Access to digital learning was also a significant predictor (OR = 2.43, p = 0.024), indicating that students with digital access were more than twice as likely to report favorable outcomes. Additionally, the use of **learner-centered teaching strategies** was significantly associated with improved outcomes (OR = 2.58, p = 0.015).

These findings highlight the combined importance of technological access, institutional capacity, and pedagogical approaches in shaping educational success

## Discussion

Nursing in Mosul City operates within a uniquely challenging yet evolving healthcare environment shaped by the long-term effects of conflict and ongoing system recovery. Despite significant damage to health infrastructure and workforce shortages in previous years, the nursing sector has demonstrated notable resilience and commitment to patient care. Nurses in Mosul often work under resource-constrained conditions, facing high patient loads, limited access to advanced technologies, and gaps in continuous professional development(10, 11). At the same time, there is a growing emphasis on improving nursing education, integrating modern clinical practices, and adopting digital learning approaches to enhance competencies. These efforts reflect a broader transition toward rebuilding a more sustainable, efficient, and equitable healthcare system, with nurses playing a central role in both service delivery and system transformation(12-14)

This study provides critical insights into the role of innovation in nursing education within a conflict-affected setting, offering both empirical evidence and strategic implications for educational reform. The findings reveal a paradoxical situation: despite high levels of student motivation and positive attitudes toward innovative learning approaches, systemic and structural barriers continue to constrain their effective implementation. This tension reflects broader challenges observed in fragile health and education systems, where the demand for transformation is high but institutional capacity remains limited. The results therefore reinforce the growing global consensus that reimagining nursing education is essential for strengthening health systems, particularly in contexts characterized by instability and resource constraints.

One of the most important findings of this study is the significant association between access to digital learning and improved student engagement and perceived clinical readiness. Students exposed to blended and technology-enhanced learning reported higher levels of confidence, critical thinking, and preparedness for clinical practice. These findings are consistent with emerging global evidence demonstrating that digital and simulation-based learning can enhance clinical reasoning and support the development of higher-order cognitive skills(15-18). In conflict-affected settings, where traditional clinical exposure may be disrupted, digital learning serves not only as a supplementary tool but as a critical mechanism for ensuring continuity of education. However, the uneven distribution of digital resources observed in this study highlights a key concern: without equitable access, digital transformation may inadvertently reinforce existing disparities rather than mitigate them.

The identification of substantial barriers including limited internet access, lack of appropriate devices, inadequate faculty training, and weak institutional support—underscores the complexity of implementing innovation in fragile contexts. Among these, faculty preparedness emerged as a particularly critical issue. While technological infrastructure is essential, the effectiveness of educational innovation ultimately depends on educators’ ability to integrate new tools and pedagogies into meaningful learning experiences. This finding emphasizes the need to shift the focus from technology adoption alone to a more comprehensive capacity-building approach that includes continuous professional development, pedagogical training, and institutional support for educators.

Institutional support was identified as the strongest predictor of positive educational outcomes, highlighting the central role of organizational leadership in driving educational transformation. Institutions that foster supportive learning environments, invest in digital infrastructure, and promote learner-centered teaching strategies are more likely to achieve meaningful improvements in student outcomes. This finding is particularly relevant in conflict-affected settings, where limited resources necessitate strategic prioritization and effective governance. Strong institutional leadership can facilitate the alignment of educational innovation with broader health system goals, ensuring that reforms are both sustainable and contextually appropriate.

From a policy perspective, the findings underscore the urgent need to position nursing education as a strategic priority within health system strengthening efforts. Investment in nursing education should not be viewed solely as an academic concern but as a critical component of workforce development and health system resilience. Policies that support digital infrastructure, equitable access to learning resources, and faculty capacity building are essential to ensure that innovation translates into tangible improvements in healthcare delivery. In countries such as Iraq, where health systems are undergoing recovery and reform, integrating educational innovation into national health and education strategies can contribute significantly to long-term system stability and performance.

Beyond policy implications, this study highlights the important role of educational innovation in shaping future nursing leadership. Educational environments that promote critical thinking, adaptability, and problem-solving equip students with the competencies required to navigate complex and rapidly changing healthcare systems. In conflict-affected settings, nurses often assume expanded roles, including leadership in crisis response and community health initiatives. Strengthening nursing education, therefore, contributes not only to individual competence but also to the development of a workforce capable of driving system-level change and innovation.

The findings also bring into focus the critical relationship between educational innovation and health equity. Disparities in access to digital tools, learning opportunities, and institutional support reflect broader structural inequities within both the education and health sectors. If not addressed, these disparities may lead to unequal skill development among graduates, ultimately affecting the quality and equity of healthcare services. Therefore, efforts to innovate nursing education must be accompanied by deliberate equity-focused strategies, including inclusive policies, targeted resource allocation, and support mechanisms for disadvantaged student groups. Ensuring equitable access to innovative educational approaches is essential to achieving broader goals of health equity and social justice.

## Limitations

Several limitations should be considered when interpreting the findings of this study. Although a multi-institutional approach was adopted to enhance representativeness, the use of a non-probability sampling strategy may still limit the generalizability of the results beyond the included universities in the Nineveh Governorate, particularly to settings with different socio-political or educational contexts.

Second, the study relied on self-reported measures of key outcomes, such as engagement, satisfaction, and perceived clinical readiness. While these indicators are commonly used in educational research and provide valuable insight into students’ perceptions, they may be subject to response bias, including overestimation or underestimation of competencies. Future studies should incorporate objective measures, such as performance-based assessments, to strengthen validity.

Third, the cross-sectional design limits the ability to establish causal relationships between variables. Although significant associations were identified, these findings should be interpreted as correlational. Longitudinal or experimental designs are recommended in future research to better examine causal pathways and the sustained impact of educational innovations.

Additionally, the study did not account for potential clustering effects at the institutional or classroom level, which may influence student experiences and outcomes. Future research using multilevel modeling approaches could provide a more nuanced understanding of these contextual influences.

Despite these limitations, the study benefits from a large sample size (n = 1100) and inclusion of multiple institutions, which enhance the robustness and contextual relevance of the findings. It contributes important empirical evidence to the limited literature on nursing education innovation in conflict-affected settings and provides a foundation for future research and policy development.

## Data Availability

All data produced in the present study are available upon reasonable request to the authors

